# Optimizing an LLM-Based Clinical Data Querying System Using Metadata Enrichment and Task Decomposition

**DOI:** 10.64898/2025.12.22.25342863

**Authors:** Weixin Liu, Bowen Qu, Pratheek Mallya, Jingyuan Wu, Kathie Thomas, Jennifer L. Hall, Juan Zhao, Zhijun Yin

## Abstract

Accessing complex clinical registries traditionally requires SQL programming expertise, limiting data accessibility for non-technical researchers. In this paper, we designed and evaluated whether a text-to-SQL solution based on large language models (LLMs) could enable natural language querying of a real-world clinical registry under strict privacy and security constraints. Using self-hosted, open-source LLMs, we developed a multi-layered optimization framework incorporating metadata enrichment, query decomposition, hybrid retrieval, and SQL self-correction. We assessed its performance across 600 queries spanning one-, two-, and three-field complexity using execution-based validation. Accuracy was improved from 88.0% to 94.5% for one-field queries and from 10.0% to 82.0% for three-field queries. Real-world testing by data scientists revealed domain-specific challenges related to coded variables, clinical ambiguity, and multi-step reasoning. We summarize key technical and operational lessons learned and discuss implications for safe, scalable deployment of LLM-assisted analytic tools in clinical registry environments.

## 1 Introduction

Clinical registries are essential for quality improvement, population surveillance, and evidence generation. The American Heart Association’s (AHA) Get With The Guidelines–Heart Failure (GWTG-HF) registry, with more than

2.44 million encounters from over 1355 hospitals, represents one of the most comprehensive national cardiovascular datasets in the United States ^1^. Although these registries are structured as tabulated data and supported by meta-data describing each data element, extracting insights typically requires SQL expertise and detailed familiarity with registry-specific variable definitions. This may create a barrier for many clinicians, researchers, and operational users who do not have programming skills and experience.

Currently, to understand the registry dataset or to generate a clinical hypothesis, researchers must submit formal data requests to centralized informatics teams, often resulting in days to weeks of latency for hypothesis testing ^2^. This centralized workflow not only delays evidence generation but also limits the ability of clinicians to perform exploratory, iterative analysis needed for quality improvement. This technical barrier creates a significant “analyst bottleneck.” Therefore, Democratizing data access through natural language interfaces could fundamentally shift this paradigm from “request-and-wait” to “self-service” analytics, accelerating the cycle of clinical discovery.

Recent advances in Large Language Models (LLMs) and Table Question Answering (Table QA) demonstrate the potential to translate natural-language questions into executable SQL queries, with increasing applications across medical domains ^3–5^. However, state-of-the-art methods developed on general-domain benchmarks like Spider ^6^ or BIRD ^7^ often fail to generalize to real-world clinical repositories. Unlike open-domain databases where column names are semantically descriptive (e.g., `customer_name`), clinical registries often utilize opaque variable codes (e.g., `HF_ETIOLOGY_1`) that require deep domain knowledge to interpret. Standard schema-linking techniques, which rely on simple string matching, are insufficient for resolving these semantic discrepancies. Furthermore, while commercial LLMs have shown high proficiency in coding tasks, relying on external APIs introduces unacceptable risks regarding data sovereignty and patient privacy, necessitating the development of capable, self-hosted alternatives.

In this study, we present an LLM-driven Natural Language to SQL (NL-to-SQL) pipeline that translates user questions into executable SQL queries for the GWTG-HF registry. Our approach integrates multiple layers, which includes metadata enrichment and query decomposition to improve SQL translation accuracy. Beyond reporting system performance, we also conducted real-world testing with AHA analysts. The key lessons learned were summarized, which are essential for safely and effectively applying LLMs to clinical registry environments.

## 2 Methods

### 2.1 Data Source and Privacy

We used the data from the GWTG-HF registry between January 1, 2005 and December 31st 2024. Patients admitted to fully participating GWTG-HF hospitals with a primary diagnosis of HF between 2005 and 2024 were included. The design of the GWTG-HF program has been previously reported ^8,9^. In brief, the GWTG-HF program is an ongoing quality improvement initiative supported by the American Heart Association and includes data from patients hospitalized with HF at participating hospitals across the US. Data is collected through manual chart abstraction and includes patient demographics, vital signs, laboratory values, prescribed medications, and in-hospital outcomes. Participating sites received approval to enroll cases without individual patient consent under the common rule or a waiver of authorization and exemption from subsequent review by their institutional review board. This study was deemed exempt from institutional review board oversight by the institutional review board for the American Heart Association. IQVIA (Parsippany, New Jersey) is the data collection and coordinating center for this study. The data can be accessed from a secure cloud platform, The American Heart Association’s Precision Medicine Platform (https://pmp.heart.org). All models were self-hosted within AHA’s secure computing environment.

### 2.2 System Architecture Overview

We developed a modular pipeline architecture (Figure 1) to enable natural language querying of the GWTG-HF registry. The system comprises six integrated components: (1) **User Interface (Streamlit)**: Web-based interface for query input, real-time status monitoring, dual SQL display (raw/corrected), field diagnostics, and query history management; (2) **Query Decomposition**: Semantic parsing module that breaks complex multi-field queries into atomic sub-queries for independent processing; (3) **Hybrid Retrieval Engine**: Combines semantic search via Facebook AI Similarity Search (FAISS) with keyword and field-name matching against the enriched metadata knowledge base; (4) **LLM Inference (Qwen2-72B)**: Self-hosted language model performing zero-shot SQL generation with retrieved context; (5) **SQL Self-Correction**: Three-stage validation pipeline (syntax fixing, field validation, business rule enforcement) ensuring query safety before execution; (6) **Clinical Database (AWS Athena)**: Serverless query engine executing validated SQL against the GWTG-HF registry and returning results as data frames.

**Figure 1.**
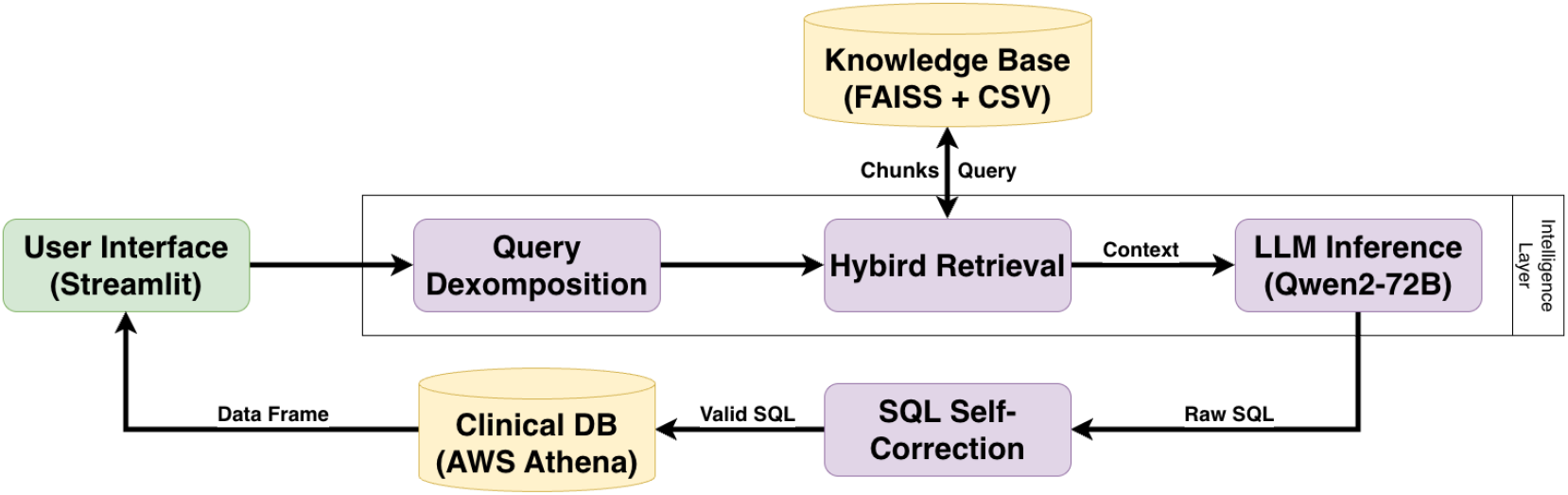
Overall System Architecture. User queries are decomposed, enhanced with retrieved context via hybrid retrieval, and used by the LLM to generate SQL. A self-correction module validates the SQL before execution against the clinical database.

The system operates through a sequential pipeline: user queries are first decomposed into atomic concepts, then each concept undergoes independent metadata retrieval. The LLM synthesizes retrieved context into executable SQL, which passes through validation layers before database execution. This architecture addresses the unique challenges of clinical data querying while maintaining strict privacy constraints through self-hosted infrastructure.

### 2.3 Preliminary Error Analysis

To systematically identify why standard Retrieval-Augmented Generation (RAG) approaches fail in real-world clinical data, we first assessed a baseline system using a **Diagnostic Subset (***n* = 200). This subset, intentionally skewed towards single-concept queries, allowed us to isolate fundamental retrieval mechanics from complex reasoning failures.

Performance was assessed using **execution-based accuracy**: a generated SQL query was considered correct only if it executed against the database without error and returned the exact same result set as a human-verified gold standard query. We established a baseline using the **Qwen2-72B model** configured with standard **“Top-5 retrieval”** (fetching the five most semantically similar metadata chunks based on vector embedding similarity). This baseline configuration achieved remarkably low accuracy on complex queries in preliminary tests, yielding distinct failure patterns. Through iterative manual analysis, we categorized these failures into three distinct types, which directly informed our optimization strategy:

- **Type 1: Retrieval Failures (21.7% of errors)**. The RAG system failed to retrieve necessary metadata columns, acting as a bottleneck even for simple queries. Patterns included: (a) *Missing enumerated values*: Questions about “Southern region patients” failed to retrieve columns containing REGION code mappings; (b) *Missing usage instructions*: The correct field (e.g., DSCHSTATI) was retrieved, but without the critical description specifying “use 6.0 for mortality calculations.”
- **Type 2: LLM Logic Errors (60.9% of errors)**. Despite the simpler query nature, the LLM still exhibited reasoning gaps: (a) *Column confusion*: Using DSCHSTATI instead of the similar DSCHOTHFACI; (b) *Redundant filtering*: Adding unnecessary WHERE clauses that over-constrain results; (c) *Enum mishandling*: Treating categorical codes as continuous values (e.g., `WHERE LVEF >= 50`instead of `WHERE LVEF = 3.0`).
- **Type 3: SQL Syntax Errors (17.4% of errors)**. The LLM produced syntactically invalid SQL, including: (a) Missing aggregation functions (`SELECT COUNT`without `(*)`); (b) Hallucinated columns (inventing `PATIENT_ID`which does not exist); and (c) Malformed BETWEEN clauses.

### 2.4 Algorithmic Optimization Framework

To address the identified failure modes, we developed a multi-layered optimization framework that integrates metadata enrichment, intelligent query decomposition, and hybrid retrieval strategies. We selected the **Qwen2-72B** as our experimental model, chosen explicitly for its superior performance on coding-specific benchmarks relative to other open-source families available at the time of study design.

#### 2.4.1 Metadata Enrichment

As the foundational optimization strategy, we first implemented metadata enrichment to systematically tackle retrieval failures (Type 1). This approach programmatically generates three complementary chunk formats for each column field: standard detailed chunks containing comprehensive specifications; concept-dense summary chunks for improved embedding retrieval; and code-specific chunks for critical enumerated values (e.g., “DSCHSTATI Code 6.0: Expired/Died”). Notably, we employed a **Human-in-the-Loop (HITL)** approach to hand-curate concept mappings for 11 critical fields with high clinical ambiguity, while applying automated extraction for the remainder. All 442 generated chunks were embedded using sentence-transformers ^10^ and indexed in FAISS ^11^.

We systematically investigated retrieval depth optimization—defined as the number of metadata chunks (*k*) fetched from the vector database—by exploring Top-*k* ∈ {5, 10, 15, 20, 25} across all query complexity levels to identify the optimal context provision strategy.

#### 2.4.2 Intelligent Query Decomposition

Even with enriched metadata, complex multi-field queries exhibited a performance ceiling due to the limitations of flat RAG architectures, where the semantic embedding of a composite query often fails to match individual column descriptions. To overcome this architectural constraint, we implemented **Intelligent Query Decomposition** as a critical optimization ^12^. The system analyzes input queries to identify constituent semantic concepts through fewshot prompting of the LLM ^13^. For example, the query “mortality rate for diabetic patients in teaching hospitals” is decomposed into three sub-concepts: [“mortality rate”, “diabetic patients”, “teaching hospitals”]. Each sub-concept undergoes independent Top-10 retrieval, with results merged alongside the original query’s retrieval results. This ensures comprehensive field coverage even when the composite query’s semantic embedding matches poorly, directly addressing the most challenging logic failures (Type 2) in our error taxonomy.

### 2.5 Engineering Enhancements

Building upon the algorithmic foundation, we integrated three advanced engineering components to bridge the gap between algorithmic capability and production reliability, informed by issues identified through independent internal testing.

#### 2.5.1 Weighted Hybrid Retrieval

To maximize retrieval precision and recall simultaneously, we implemented a **weighted hybrid retrieval system** combining three complementary scoring mechanisms: *Semantic Similarity* (*w* = 0.3) uses FAISS-based dense retrieval to capture conceptual relationships; *Keyword/Alias Matching* (*w* = 0.4) matches queries against curated synonym lists to handle vocabulary gaps; and *Field Name Exact Matching* (*w* = 0.3) ensures direct mentions (e.g., “LVEF”) are never missed. We optimized these weights via grid search on the validation tuning set, with chunks ranked by combined score for top-20 selection.

#### 2.5.2 SQL Self-Correction Pipeline

To address residual syntax and logic errors in generated SQL, we implemented a multi-pass **SQL self-correction pipeline** with three specialized modules ^12,14^. The *Syntax Correction* module repairs common errors like malformed BETWEEN clauses and table name quoting. The *Field Validation* module checks all extracted column names against a whitelist of retrieved fields to prevent hallucinations. Finally, the *Business Rule Correction* module enforces domain-specific logic, such as ensuring correct mortality codes (DSCHSTATI=6.0) and proper denominator handling for proportion calculations. The system iterates up to 2 passes, sequentially applying corrections until stability is reached.

#### 2.5.3 Enhanced Few-Shot SQL Generation

To further improve LLM generation quality, we implemented **enhanced SQL generation via Few-Shot Injection** by augmenting prompts with relevant SQL examples extracted from the enriched CSV metadata dictionary. For each field retrieved during RAG, the system checks for associated `SQL_Examples`(e.g., “Calculate mortality rate: `SELECT…`“) and injects them as few-shot demonstrations. This significantly improves adherence to medical coding conventions and reduces encoding errors.

### 2.6 Evaluation Methodology

#### 2.6.1 Dataset Construction and Zero-Shot Evaluation Design

Unlike traditional supervised learning systems requiring train-validation-test splits to prevent overfitting, our system performs **zero-shot inference**—defined here as generating SQL utilizing the model’s inherent capabilities without updating its internal weights on our specific dataset. We utilized a frozen version of **Qwen2-72B**, which was pretrained on massive general corpora of text and code. This fundamental architectural difference eliminates the risk of test set contamination, allowing methodologically sound reuse of the same test dataset across multiple configuration experiments ^13^.

The evaluation question distribution was anchored on representative clinical questions curated by the American Heart Association (AHA) data science team. The GPT-4o model was used primarily for semantic rewriting, controlled diversification, and compositional expansion of these seed questions across 1-, 2-, and 3-field complexities. Crucially, no patient data or private schema values were exposed to the GPT-4o API during this process; the model operated solely on generic semantic templates without independently generating or simulating clinical needs:

- **Validation Pool (***n* = 400): A held-out repository used strictly for system diagnostics and hyperparameter selection, divided into two purpose-specific subsets:

- *Subset A: Diagnostic Set (n* = 200): Predominantly single-field queries used for the error analysis in Section 2.3 to identify fundamental retrieval and syntax failure modes.
- *Subset B: Hyperparameter Selection Set (n* = 200): A stratified set (balanced across 1, 2, and 3-field complexities) used for grid search operations, specifically for selecting retrieval depth (*k*) and optimizing hybrid retrieval weights.

- **Test Set (600 queries):** The primary evaluation corpus for final performance reporting, completely exclusive from the validation pool:

- 1-field queries (*n* = 200): Single-concept questions
- 2-field queries (*n* = 200): Questions requiring two fields
- 3-field queries (*n* = 200): Questions requiring three fields

#### 2.6.2 Methodological Validity and Evaluation Metrics

All experiments use frozen pre-trained LLMs without gradient-based parameter updates. Therefore our procedure constitutes configuration selection (e.g., retrieval weights, Top-k) rather than model training. The use of Subset B ensures that hyperparameters are optimized for **stratified difficulty levels**, while the Test Set remains unseen, consistent with standard zero-shot evaluation protocols ^13^.

Accuracy was measured using **Exact Match (EM)**: generated SQL is correct if and only if it produces identical query results to gold-standard SQL when executed on the actual GWTG-HF database. This metric is more stringent than syntactic correctness but better reflects real-world utility. All SQL generation used a temperature of 0.2 to minimize randomness and favor deterministic outputs.

## 3 Results

All results reported in this section are obtained on the held-out 600-query test set described in Section 2.6. Optimization strategies were selected based on validation-set analysis (Section 2.3). We present the performance improvements hierarchically, starting from the standard RAG baseline and demonstrating the systematic accuracy gains achieved through algorithmic optimizations and engineering enhancements.

### 3.1 Baseline Performance

We established a baseline using standard (non-enriched) metadata retrieval with the Qwen2-72B model at Top-*k* = 10 configuration, determined via validation set analysis. The baseline achieved 88.0% accuracy on simple 1-field queries, but performance degraded sharply for complex queries: 32.0% for 2-field queries and only 10.0% for 3-field queries. This confirms the severe performance ceiling of standard RAG for multi-field reasoning in clinical domains.

### 3.2 Impact of Metadata Enrichment

Table 1 benchmarks the impact of metadata enrichment against the standard RAG baseline. For Qwen2-72B, the gains were pronounced: 2-field queries improved from 32.0% (baseline with raw metadata, *k* = 10) to 61.0% with enriched metadata (*k* = 10), representing a +29.0 percentage point improvement. Similarly, 3-field queries improved from 10.0% (baseline, *k* = 10) to 33.0% (enriched, *k* = 10), a +23.0 point gain. This universal improvement pattern proves metadata enrichment as a foundational requirement. Notably, improvements are most pronounced for mid-to-high complexity queries (2-field and 3-field), where retrieval quality critically determines success. The enriched metadata ensures that critical information, such as enumerated value codes, usage instructions, and semantic synonyms, is reliably retrieved, directly addressing the Type 1 retrieval failures identified in our error analysis.

**Table 1:** Performance Improvement from Metadata Enrichment with Qwen2-72B (600-Query Test Set).

| Query Type | Top-k | Standard RAG (%) | Enriched (%) | Gain |
| --- | --- | --- | --- | --- |
| 1-field | 5 | 87.0 | 92.0 | +5.0 |
|  | 10 | 88.0 | 92.0 | +4.0 |
| 2-fields | 5 | 29.0 | 46.5 | +17.5 |
|  | 10 | 32.0 | 61.0 | +29.0 |
| 3-fields | 5 | 3.0 | 5.5 | +2.5 |
|  | 10 | 10.0 | 33.0 | +23.0 |

### 3.3 Impact of Query Decomposition

While metadata enrichment provided substantial improvements, complex multi-field queries still exhibited a performance ceiling (61.0% for 2-field, 33.0% for 3-field with Qwen2-72B). This ceiling stems from a fundamental limitation of flat RAG: when a query mentions multiple concepts, the semantic embedding of the composite query may poorly match individual field descriptions.

To overcome this limitation, we implemented intelligent query decomposition. Table 2 demonstrates substantial improvements: Qwen2-72B on 2-field queries improved from 61.0% (enriched only) to 89.0% (enriched + decomposition), representing a +28.0 percentage point gain. For 3-field queries, accuracy leaped from 33.0% to 77.5% (+44.5 points)—more than doubling the accuracy and breaking through the flat RAG performance ceiling. These results confirm that advanced RAG strategies yield transformative benefits when paired with capable reasoning engines, effectively resolving the semantic dilution problem in complex clinical queries.

**Table 2:** Query Decomposition Performance Gains with Qwen2-72B (600-Query Test Set)

| Query Type | Top-k | Enriched Only (%) | +Decomposition (%) | Gain |
| --- | --- | --- | --- | --- |
| 1-field | 10 | 92.0 | 92.0 | 0.0 |
| 2-fields | 10 | 61.0 | **89.0** | +28.0 |
|  | 20 | 72.5 | 87.5 | +15.0 |
| 3-fields | 20 | 33.0 | **77.5** | +44.5 |
|  | 25 | 38.5 | 75.5 | +37.0 |

### 3.4 Algorithmic Ablation Study

To isolate the individual contributions of each algorithmic optimization strategy, we conducted a comprehensive ablation analysis using Qwen2-72B, as detailed in Table 3.

**Table 3:** Ablation Study: Algorithmic Component-wise Contributions (Qwen2-72B, 600-Query Test Set)

| Configuration | 1-Field (%) | 2-Field (%) | 3-Field (%) |
| --- | --- | --- | --- |
| (1) Baseline (Raw Metadata) | 88.0 | 32.0 | 10.0 |
| (2) + Metadata Enrichment | 92.0 | 61.0 | 33.0 |
| (3) + Enrichment + Decomposition (Full) | **92.0** | **89.0** | **77.5** |
| *Incremental Gains:* |  |  |  |
| Enrichment over Baseline | +4.0 | +29.0 | +23.0 |
| Decomposition over Enriched | 0.0 | +28.0 | +44.5 |
| **Total Algorithmic Gain** | **+4.0** | **+57.0** | **+67.5** |

We first established the **baseline performance** (Row 1) using standard (non-enriched) metadata retrieval at a Top-k configuration of *k* = 10, determined via validation tuning set analysis. While the system achieved 88.0% accuracy on simple 1-field queries, performance degraded sharply for complex queries: 32.0% for 2-field queries and only 10.0% for 3-field queries. This confirms the severe performance ceiling of standard RAG for multi-field reasoning.

Upon this baseline, **metadata enrichment** (Row 2) elevated performance to 33.0% on 3-field queries (+23.0 points), primarily addressing retrieval failures. Finally, **intelligent query decomposition** (Row 3) propelled accuracy to 77.5% (+44.5 points over the enriched baseline), breaking through the flat RAG ceiling by ensuring comprehensive field coverage. This clearly demonstrates that both strategies are necessary: enrichment establishes a foundational improvement, while decomposition yields marginal gains that scale dramatically with query complexity.

### 3.5 Final Production System Performance

Integrating all engineering enhancements (weighted hybrid retrieval, SQL self-correction pipeline, and few-shot injection) with the algorithmic baseline yields the final production system performance. As shown in Table 4, the fully integrated system achieves 94.5% accuracy on 1-field queries, 92.5% on 2-field queries, and 82.0% on 3-field queries.

**Table 4:** Final Production System Performance (600-Query Test Set)

| Configuration | 1-Field (%) | 2-Field (%) | 3-Field (%) |
| --- | --- | --- | --- |
| Algorithmic Baseline<br>(Enrichment + Decomposition) | 92.0 | 89.0 | 77.5 |
| **Full Production System**<br>(+ All Engineering Enhancements) | **94.5** | **92.5** | **82.0** |
| *Engineering Contribution:* |  |  |  |
| Total Improvement | +2.5 | +3.5 | +4.5 |

The engineering layers contribute a cumulative gain of +4.5 percentage points on complex 3-field queries (from 77.5% algorithmic baseline to 82.0% final system), validating their necessity for production robustness. For 2-field queries, the production system achieves 92.5% (up from 89.0% baseline, +3.5 points), and for 1-field queries, 94.5% (up from 92.0%, +2.5 points). These incremental but meaningful improvements demonstrate that holistic engineering integration—combining hybrid retrieval, self-correction, and few-shot learning—is essential to bridge the gap between algorithmic capability and production-ready reliability.

## 4 Discussion

### 4.1 Principal Findings and Implications

This research supports a hierarchical, error-driven methodology for constructing deployment-focused Text-to-SQL systems under real-world constraints. Our preliminary error taxonomy (21.7% retrieval failures, 78.3% generation failures) guided targeted optimizations yielding substantial gains. This two-phase improvement supports our central hypothesis: practical deployment-ready systems require both algorithmic innovation and engineering robustness.

The most significant algorithmic finding is the synergistic interaction between model capability and decomposition strategy. Qwen2-72B achieved a 44.5 point improvement on 3-field queries (from 33.0% to 77.5%), demonstrating that advanced RAG strategies do not merely compensate for weak models, but rather enable capable models to leverage their reasoning capacity effectively.

The deployed hybrid retrieval system addresses a fundamental limitation of pure semantic retrieval in medical domains: terminology exhibits both high synonymy (many terms for “mortality”) and high homonymy (LVEF refers to both the concept and the database field). By combining semantic, keyword, and exact-match signals, the system achieves both high recall (via semantic similarity) and high precision (via explicit matching).

The SQL self-correction system mitigates a critical operational risk: deploying LLM-generated SQL directly to production databases risks data corruption or security breaches from malformed queries. Our three-module correction pipeline (syntax, field validation, business rules) establishes a safety layer between LLM output and database execution. This addresses a gap in existing Text-to-SQL research, which typically evaluates syntactic correctness but not semantic validity against domain-specific business logic.

### 4.2 Comparison to Related Work and Benchmarks

Contemporary Text-to-SQL research focuses on large benchmarks (Spider, BIRD) using commercial API access (GPT-4, Claude). Our work differs fundamentally: strict privacy constraints mandate self-hosted deployment, complex medical coding requires domain-specific optimization, and production deployment demands real-time performance and robustness. While we cannot directly compare to Spider SOTA (which assumes GPT-4 API access), our 89.0% accuracy on 2-field clinical queries is qualitatively competitive with reported DIN-SQL and DAIL-SQL performance on similarly complex BIRD queries ^12,15^, though direct comparison is infeasible due to dataset differences (GWTG-HF vs. BIRD schema complexity, privacy constraints precluding BIRD evaluation).

Our query decomposition strategy shares conceptual similarity with DAIL-SQL’s demonstration selection and DIN-SQL’s schema linking ^12,15^, but operates at the semantic query level rather than schema level. Future work could explore combining these complementary approaches. Our hybrid retrieval extends recent work on dense-sparse retrieval fusion in information retrieval ^16^, applying similar principles to the Text-to-SQL domain.

### 4.3 Limitations and Future Directions

Several limitations warrant acknowledgment. First, our evaluation focuses on structured data and does not include unstructured text (e.g., clinical notes). While metadata enrichment and decomposition are domain-agnostic, generalization to other schemas (MIMIC-III ^17^, PCORnet, Epic Clarity) requires adaptation and validation. Second, we focus on queries with different levels of complexity generated from seed questions. Although this approach ensures coverage across complexity levels, the resulting questions may differ from original clinician’s real-world queries in vocabulary, ambiguity, and real-world information needs. Internal evaluation is still undergoing, and future work should include prospective user studies with clinical researchers to evaluate performance on real-world questions and identify additional failure modes. Third, the system handles only SELECT queries; future versions should support analytical patterns (subqueries, window functions, CTEs).

Another interesting future direction is adaptive strategy selection. Our results show decomposition benefits complex queries. However, it can exhibit diminishing returns on atomic, single-concept queries. An intelligent router using a lightweight BERT classifier (trained on query features: token count, entity count, syntactic depth) could automatically select standard RAG versus decomposition pathways, maximizing both accuracy and efficiency ^18^. Additional ongoing work includes uncertainty quantification to flag low-confidence generations ^19^ and multi-turn dialog to have end user in the generation loop for resolving query disambiguation, which is not uncommon in practice ^12,20^.

## 5 Conclusion

This study shows that building reliable Text-to-SQL systems for specialized clinical databases requires a structured, multi-layered approach grounded in systematic error analysis. A key finding is that neither LLM scale nor RAG sophistication alone is enough: strong performance emerges from their coordinated use, guided by an understanding of retrieval and generation failure modes. As healthcare organizations seek to broaden access to clinical data, the methods validated here—error-driven optimization, hybrid retrieval, SQL self-correction, and configuration-driven design—provide a practical and reproducible path forward that maintains strict privacy and accuracy standards. The two-phase development strategy, combining algorithmic improvements with focused engineering enhancements, offers a useful framework for building Text-to-SQL systems in privacy-sensitive settings. Overall, this work highlights that effective clinical data querying depends not on model size alone, but on carefully engineered systems tailored to real-world constraints and failure modes.

## Data Availability

The data can be accessed from a secure cloud platform, The American Heart Association's Precision Medicine Platform (https://pmp.heart.org). Access is subject to approval by the American Heart Association.

## Acknowledgments

This project is supported by the Doris Duke Foundation (grant no. 25DECCAGWTG1372664).

## Notes

### Competing Interest Statement

J.L.H., K.T., J.Z., P.M., and J.W. are employees of the American Heart Association. Other authors declare no competing interests.

### Author Declarations

The Institutional Review Board of the American Heart Association waived ethical approval for this work.

